# Sex hormone and metabolic dysregulations are associated with critical illness in male Covid-19 patients

**DOI:** 10.1101/2020.05.07.20073817

**Authors:** Maria Schroeder, Berfin Schaumburg, Zacharias Müller, Ann Parplys, Dominik Jarczak, Axel Nierhaus, Andreas Kloetgen, Bettina Schneider, Manuela Peschka, Fabian Stoll, Tian Bai, Henning Jacobsen, Martin Zickler, Stephanie Stanelle-Bertram, Geraldine de Heer, Thomas Renné, Andreas Meinhardt, Joerg Heeren, Jens Aberle, Alice C. McHardy, Hartmut Schlüter, Jens Hiller, Sven Peine, Lothar Kreienbrock, Karin Klingel, Stefan Kluge, Gülsah Gabriel

**Affiliations:** Department of Intensive Care Medicine, University Medical Center Hamburg-Eppendorf, Germany; Department for Viral Zoonoses-One Health, Heinrich Pette Institute, Leibniz Institute for Experimental Virology, Hamburg, Germany; Computational Biology of Infection Research, Helmholtz Centre for Infection Research, Braunschweig, Germany; Department of Biometry, Epidemiology and Information Processing, University of Veterinary Medicine Hannover, Germany; Institut for Clinical Chemistry and Laboratory Medicine, University Medical Center Hamburg-Eppendorf, Germany; Institute of Anatomy and Cell Biology, Justus-Liebig University of Giessen, Germany; Institute for Biochemistry and Molecular Cell Biology, University Medical Center Hamburg-Eppendorf, Germany; Department of Endocrinology, Diabetology, Obesity and Lipids, University Medical Center Hamburg-Eppendorf, Germany; German Center for Infection Research (DZIF), Germany; Institute for Transfusion Medicine, University Medical Center Hamburg-Eppendorf, Germany; Institute for Pathology and Neuropathology, University Hospital Tuebingen, Germany; Institute for Virology, University for Veterinary Medicine Hannover, Germany

## Abstract

Males develop more severe SARS-CoV-2 infection related disease outcome than females. Herein, sex hormones were repeatedly proposed to play an important role in Covid-19 pathophysiology and immunity. However, it is yet unclear whether sex hormones are associated with Covid-19 outcome in males and females. In this study, we analyzed sex hormones, cytokine and chemokine responses as well as performed a large profile analysis of 600 metabolites in critically-ill male and female Covid-19 patients in comparison to healthy controls and patients with coronary heart diseases as a prime Covid-19 comorbidity. We here show that dysregulated sex hormones, IFN-γ levels and unique metabolic signatures are associated with critical illness in Covid-19 patients. Both, male and female Covid-19 patients, present elevated estradiol levels which positively correlates with IFN-γ levels. Male Covid-19 patients additionally display severe testosterone and triglyceride deficiencies as compared to female patients and healthy controls. Our results suggest that male Covid-19 patients suffer from multiple metabolic disorders, which may lead to higher risk for fatal outcome. These findings will help to understand molecular pathways involved in Covid-19 pathophysiology.

## Introduction

The current SARS-CoV-2 pandemic continues taking its toll on human health with currently 1.61 million lives lost worldwide (as of 14^th^ December 2020). SARS-CoV-2 was first reported in humans in December 2019 in Wuhan, China ^1^. On February 11^th^ 2020, the World Health Organization (WHO) named the disease caused by SARS-CoV-2, COVID-19 (coronavirus disease 2019). One month later, on 11^th^ March 2020, the WHO declared COVID-19 as a pandemic. The clinical spectrum of SARS-CoV-2 infection is broad, ranging from mild upper respiratory illnesses to severe primary pneumonia with respiratory failure, multi-organ failure and death ^2^. Retrospective cohort studies revealed risk factors associated with disease severity and death. A study from Wuhan, China, which enrolled 191 inpatients on hospital admission since its first occurrence in China in December 2019, reported that older age and comorbidities, such as hypertension, diabetes and coronary heart diseases being among the top three, present poor prognostic markers at an early stage ^2^. Another study from the UK linked 10,926 COVID-19-related deaths pseudonymously to primary care records of 17 million individuals, identifying being male, older age, diabetes, asthma, obesity as well as chronic heart diseases among the comorbidities associated with Covid-19 related death ^3^.

Thus, there is increasing evidence that being male constitutes a major risk factor associated with SARS-CoV-2 fatality. However, the underlying factors of sex disparity observed in Covid-19 remain unclear yet. We have recently shown using the golden hamster model, that SARS-CoV-2 infection attacks the reproductive organs and causes massive dysregulation of sex hormones in infected male and female animals ^4^. In the young and lean golden hamsters without comorbidities, the males had reduced plasma testosterone levels combined with elevated plasma estradiol levels, unlike females who showed reduced plasma estradiol levels upon SARS-CoV-2 infection ^4^. Thus, we wanted to study whether the observed dysregulation in sex hormones upon SARS-CoV-2 infection is also present in Covid-19 patients and poses a risk factor for disease severity.

Therefore, we herein analyzed sex hormones, cytokine responses and more than 600 metabolites in critically-ill male and female Covid-19 patients in comparison to healthy controls and patients with coronary heart diseases as one of the top comorbidities present in Covid-19 patients.

## Results

### More male patients with Covid-19 required intensive care than women

A total of *n*=136 SARS-CoV-2 PCR-positive patients were admitted to the Clinic for Intensive Care Medicine, at the University Medical Center Hamburg-Eppendorf within the period 9^th^ March to 9^th^ December 2020. Of these, *n*=88 (65%) were male and *n*=48 (35%) were female. A total of *n*=41 (30%) patients died, of which *n*=25 (61%) were male and *n*=16 (39%) were female (**Figure 1**).

**Figure 1.**
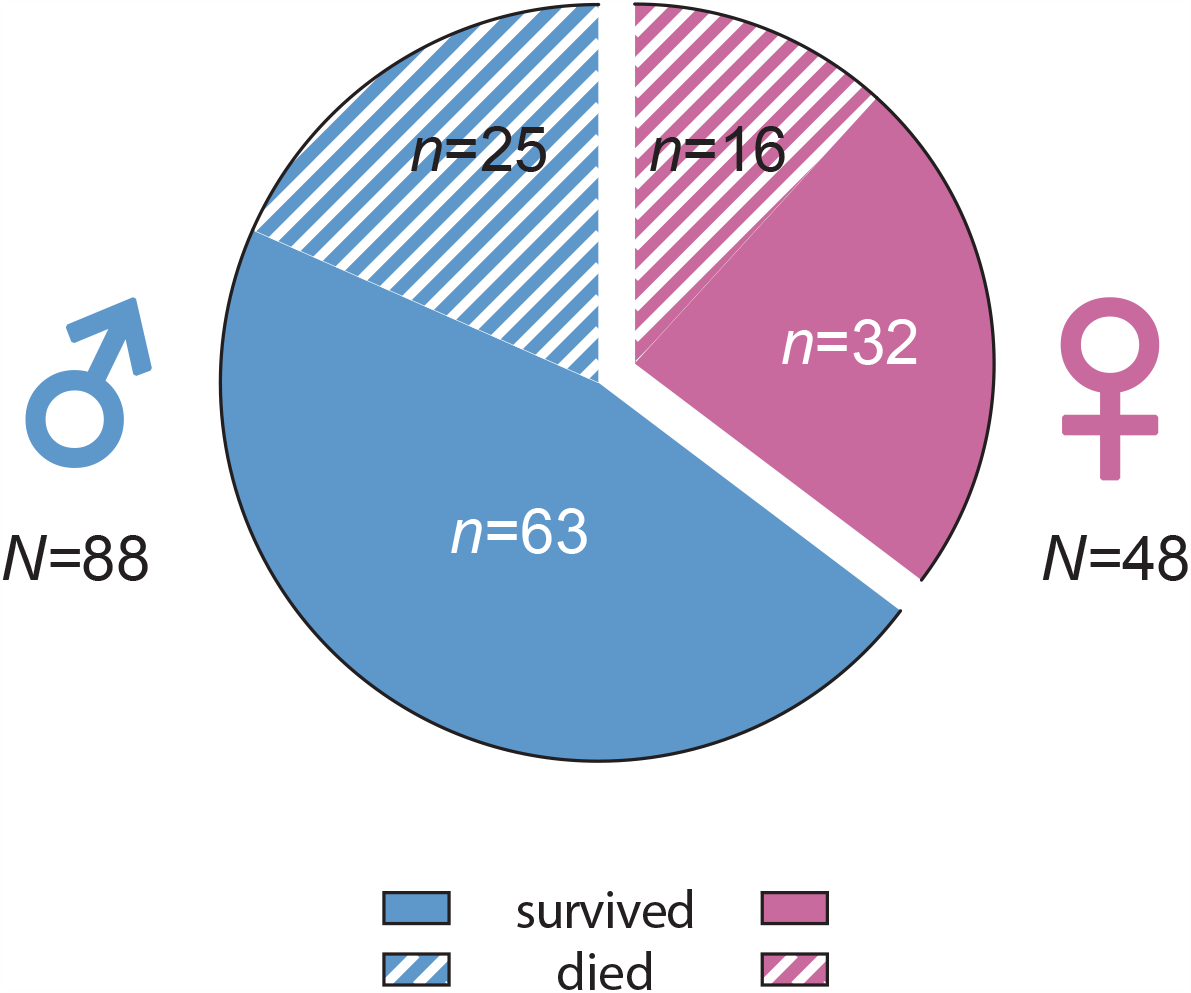
Covid-19 patients in the intensive care unit. Shown are Covid-19 patient numbers at the Clinic for Intensive Care Medicine, at the University Medical Center Hamburg-Eppendorf within the period 9^th^ March-9^th^ Decemebr 2020. Patients were further subdivided in surving (males, *n*=63; females, *n*=32) and deceased (males, *n*=25; females, *n*=16) patients.

### Severe Covid-19 in men is associated with reduced androgen and increased estrogen levels

First, we wanted to assess whether SARS-CoV-2 infection mediated alterations in sex hormone levels proposed before in an animal model ^4^ are also observed in Covid-19 patients and pose a risk factor for severe outcome. Therefore, we recruited male and female Covid-19 patients (*n*=50) who were admitted to an intensive care unit (**Table 1** and **Table S1**). The median age in Covid-19 patients was comparable between males (63 years, interquartile range (IQR) 15 years) and females (67 years, IQR 8.5 years), respectively. All patients presented at least one comorbidity with coronary heart diseases (CHD), diabetes type II and obesity being among the most frequent (**Table S1**). However, diabetes type II (*P=*1.0) and obesity (*P=*1.0) showed same frequencies in male and female Covid-19 patients. Albeit statistically not significant (*P=*0.6), more coronary heart diseases (CHD) patients were present in the male compared to the female Covid-19 cohort, which might reflect extended frequency of CHD in men ^5^. Since obesity and type II diabetes were evenly distributed between male and female Covid-19 patients, sex hormone analysis might be shifted due to this unbalance of CHD presence. Therefore, we recruited age- and sex-matched male and female SARS-CoV-2 negative CHD patients (*n*=39) as an internal control. As an additional healthy control, we recruited age-and sex-matched SARS-CoV-2 negative male and female blood donors (HC) (*n*=50) (**Table 1**). We then measured major sex hormones (testosterone, dihydrotestosterone, estradiol and estrone) in Covid-19 patients and the respective control cohorts.

**Table 1:** Patient Demographics.

| Characteristics | HC<br>(n=50) | CHD<br>(n=39) | COVID-19<br>(n=50) |
| --- | --- | --- | --- |
| **Sex, No (%)** |  |  |  |
| Male | 30 (60) | 25 (64) | 39 (78) |
| Female | 20 (40) | 14 (36) | 11 (22) |
| **Age, mean (CV), y** |  |  |  |
| Male | 58.5 (12.3) | 65.7 (16.9) | 63.0 (19.7) |
| Female | 55.7 (17) | 64.6 (21.6) | 65.8 (16.7) |

Testosterone levels were reduced in the plasma of male CHD patients compared to male HC (*P*<0.0001) (**Figure 2a**). This is in line with previous reports on reduced testosterone levels in male patients with cardiovascular diseases ^6^. Of note, plasma testosterone levels were strongly reduced in male Covid-19 patients as compared to HC and CHD cohorts (*P*<0.0001 and *P=*0.0399, respectively). Most Covid-19 males had testosterone levels far below clinical reference values ^7,8^ suggesting severe testosterone consumption and/or synthesis. In females, plasma testosterone levels were comparable within the HC, CHD and Covid-19 cohorts without significant changes (**Figure 2b**). Within the measured testosterone levels, a statistical significant interaction between sex and Covid-19 and CHD cohort appears (*P*<0.0001 and *P*<0.0163, respectively) (**Figure 2c**) further confirming testosterone deficiency in males. Testosterone is further metabolized to dihydrotestosterone by 5-α reductase. Dihydrotestosterone also acts as an androgen and plays a key role in activating the transcription of various genes and activation of various immune cells similar to testosterone ^9^. Thus, we wanted to assess whether alterations in testosterone levels detected in Covid-19 patients are also reflected in its most potent metabolite. In male Covid-19 patients, dihydrotestosterone levels were significantly reduced compared to HC males (*P*<0.0001) (**Figure 2d**). In line, a substantial proportion of plasma dihydrotestosterone levels in Covid-19 males were even below the lowest reference range, confirming dihydrotestosterone deficiency in men. In contrast, dihydrotestosterone levels were comparable between female Covid-19 and HC cohorts within clinical references (*P*=0.9568) (**Figure 2e**). Further statistical significant interaction validates dihydrotestosterone deficiency in Covid-19 males (*P*<0.0001) (**Figure 2f**).

**Figure 2.**
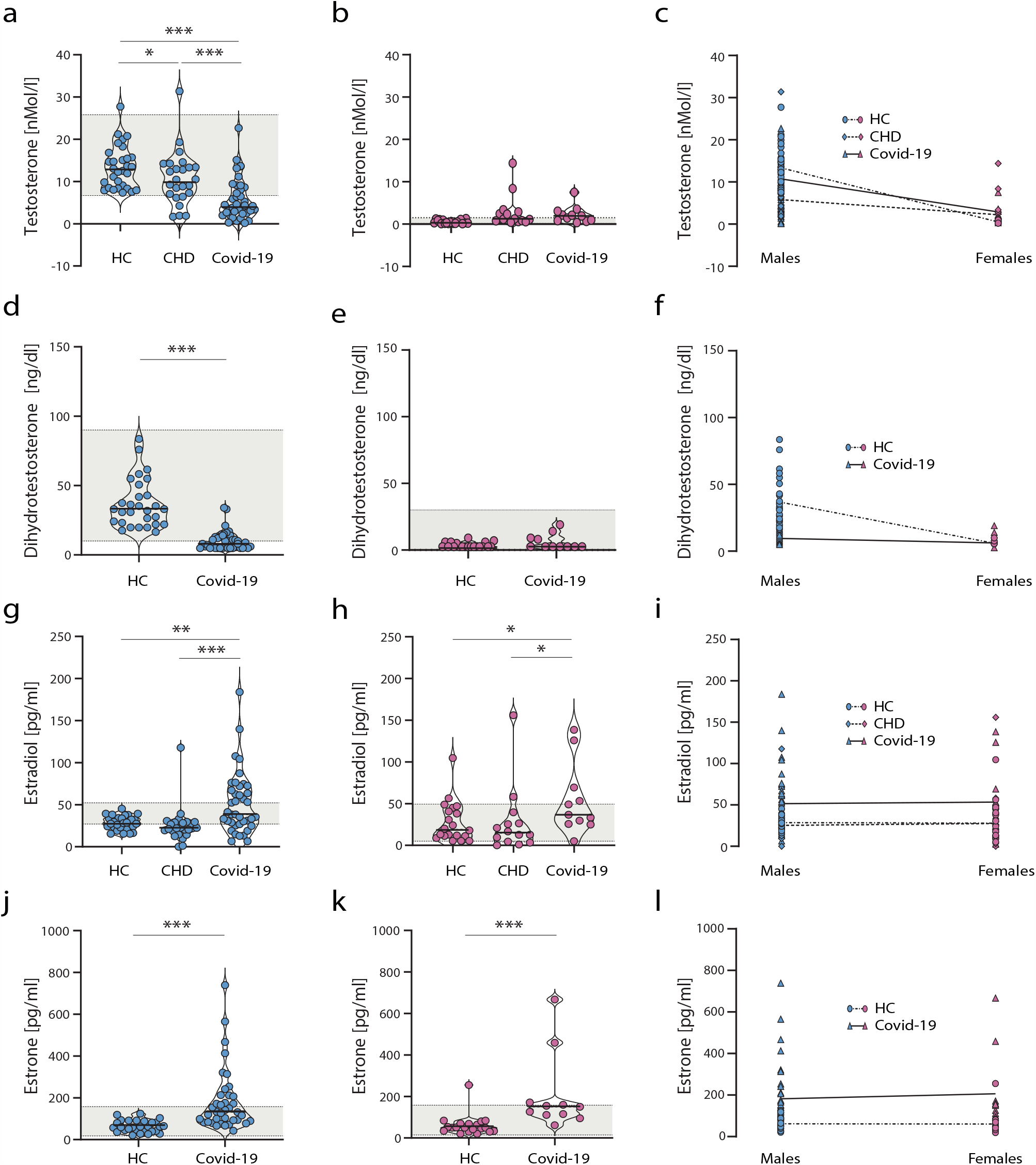
Sex hormone levels in Covid-19 patients, healthy controls and patients with coronary heart diseases. Testosterone (a-c), dihydrotestosterone (d-f), estradiol (g-i) and estrone (j-l) levels were measured in plasma obtained from COVID-19 patients, aged-matched healthy donors (HD) and coronary heart disease patients (CHD). Blue dots represent males (HD, *n*=30; CHD, *n*=25; COVID-19, *n*=30) and red dots represent females (HD, *n*=20; CHD, *n*=14; COVID-19, *n*=11) (a-l). Values are shown as median. The laboratory assessed hormone reference ranges are indicated in grey. Statistical significance was assessed via Two-Way-ANOVA.

Estradiol levels were comparable in the plasma between HC and CHD males (*P*=0.6802). However, plasma estradiol levels were significantly increased in Covid-19 male patients unlike HC and CHD cohorts (*P*=0.0017 and *P*=0.0007, respectively) (**Figure 2g**). In females, plasma estradiol levels were comparable between HC and CHD groups (*P*=0.9764). In Covid-19 females, however, plasma estradiol levels were significantly increased as compared to the HC controls and CHD patients (*P*=0.0222 and *P*=0.0309, respectively) (**Figure 2h**). These statistical significance of elevated estradiol levels in Covid-19 males and females was shown in a simultaneous analysis (**Figure 2i**). To assess whether the increase in estradiol levels is attributed to a general increase in estrogens, we next measured estrone concentrations. Estrone levels in the plasma of Covid-19 males were significantly higher compared to HC males (*P*<0.0001) (**Figure 2j**). Similarly, estrone levels were significantly elevated in the plasma of female Covid-19 patients unlike HC females (*P*=0.0009) (**Figure 2k**). There was no statistical significant interaction between sex and cohort observed further validating the sex-independent increase in estrone levels (**Figure 2l**).

Collectively, these findings show that male and female Covid-19 patients present increased estrogen levels (estradiol and estrone). However, male Covid-19 patients additionally suffer from a severe androgen (testosterone and dihydrotestosterone) deficiency.

### Male Covid-19 patients present primary and secondary hypogonadism

To shed light on the origin of severe testosterone deficiency in male COVID-19 patients observed in this study, we further analyzed related hormones (**Table 2**). Free testosterone levels were reduced in 66.7% of male Covid-19 patients compared to reference values. Conversely, 54.5% of female Covid-19 patients presented elevated levels of free testosterone. Thus, changes in total testosterone levels reported above correlate with levels of free bioavailable testosterone levels in the respective sex. We then measured levels of the sex hormone-binding globulin (SHBG) since the majority (98%) of total testosterone is bound to SHBG and only 2% is in its free, bioavailable form. Thus, in some cases, testosterone deficiencies might be masked by elevated SHBG levels. In 28.2% of male COVID-19 patients, SHBG levels were elevated, which might suggest masked testosterone deficiencies in some patients. Luteinizing hormone (LH) levels were elevated in 30.8% of male COVID-19 patients, while being within the normal range in all female patients. Interestingly, 7 out of the 28 male patients with low total testosterone levels presented elevated LH levels at the same time (data not shown), suggesting impairment of Leydig cell steroidogenesis in 25.0% of the male patients. Follicle stimulating hormone (FSH) levels were elevated in 12.8% of male patients. Elevated FSH levels in these male patients were combined with elevated LH levels. In 45.5% of female patients, FSH levels were reduced, which may indicate loss of ovarian function. This would be in line with the postmenopausal status of the 10 out of 11 Covid-19 females in our cohort. Other hormones, such as thyroid stimulating hormone (TSH) and T4 were within normal ranges in the majority of male and female patients. Cortisol levels were elevated in 56.4% of male and 81.8% of female COVID-19 patients.

**Table 2:** Hormone Levels in Covid-19 Patients.

| Hormone Levels | Covid-19 Males (n=39) | Covid-19 Females (n=11) |
| --- | --- | --- |
| **Free testosterone** |  |  |
| Normal males: 20-39 yr: 7-22.7 pg/ml | 13 (33.3%) |  |
| 40-60 yr: 6.3-17.8 pg/ml |  |  |
| ≥61 yr: 2.5-17.8 nMol/l |  |  |
| Low (all age groups, below reference) | 26 (66.7%) |  |
| Normal females 40-60 yr: ≤2.3 pg/ml |  | 5 (45.5%) |
| ≥61 yr: ≤2.1 pg/ml |  |  |
| High (all age groups, above reference) |  | 6 (54.5%) |
| **Sex hormone-binding globulin** |  |  |
| Normal males: 10-40 nMol/l | 27 (69.2%) |  |
| Low: <10 nMol/l | 1 (2.6%) |  |
| High: 41-100 nMol/l | 7 (17.9%) |  |
| Very High: ≥101 nMol/l | 4 (10.3%) |  |
| Normal females: 26-110 nMol/l |  | 7 (63.6%) |
| High: ≥110 nMol/l |  | 1 (9.1%) |
| Low: <26 nMol/l |  | 3 (27.3%) |
| **Luteinizing hormone** |  |  |
| Normal males: 0-8.6 mIU/ml | 27 (69.2%) |  |
| High: ≥8.7 mIU/ml | 12 (30.8%) |  |
| Normal females: <58.5 mIU/ml |  | 11 (100%) |
| **Follicle stimulating hormone** |  |  |
| Normal males: 1.5-12.4 mIU/ml | 32 (82.1%) |  |
| High: 12.5-25 mIU/ml | 5 (12.8%) |  |
| Low: <1.5 mIU/ml | 2 (5.1%) |  |
| Normal females: 25.8-134.8 mIU/ml |  | 6 (54.5%) |
| Low: 10-25.7 mIU/ml |  | 5 (45.5%) |
| **Thyroid stimulating hormone** |  |  |
| Normal: 0.27-4.2 μU/ml | 30 (76.9%) | 7 (63.6%) |
| High: >4.2 μU/ml | 2 (5.1%) | 1 (9.1%) |
| Low: <0.27 μU/ml | 7 (18.0%) | 3 (27.3%) |
| **Free T4** |  |  |
| Normal: 8-17 ng/dl | 38 (97.4%) | 11 (100%) |
| High: >17 ng/dl | 1 (2.6%) |  |
| **Cortisol** |  |  |
| Normal: 2.5-19.5 μg/dl | 16 (40.0 %) | 2 (18.2%) |
| High: 23-30 μg/dl | 22 (56.4%) | 9 (81.8%) |
| Low: <23 μg/dl | 1 (2.6%) |  |

These findings suggest that in 25% of the male COVID-19 patients with low total testosterone levels, testosterone deficiency is likely of testicular origin. Thus, in 75% of male patients, the origin of testosterone deficiency remains unclear.

### Disease severity in male Covid-19 patients correlates with elevated cytokine and chemokine responses

Next, we compared cytokine and chemokine patterns in male and female Covid-19 patients. Therefore, we analyzed a panel of 27 different cytokines and chemokines in the plasma of Covid-19 patients and correlated to disease severity as assessed by the Sequential Organ Failure Assessment Score (SOFA). In general, cytokine and chemokine responses increased with increasing disease severity in male and female patients with the exception of IL-12 in males (**Figure 3**). In male Covid-19 patients, particularly IFN-γ (*P*=0.0301), IL-1RA (*P*=0.0160), IL-6 (*P*=0.0145), MCP-1 (*P*=0.0052) and MIP-1α (*P*=0.0134) levels were significantly elevated in those with higher SOFA scores (8-11) compared to those with lower SOFA scores (2-3) (**Figure 3a-e**). In female patients, TNF-α levels were significantly higher in those with high SOFA scores compared to those with low SOFA scores (*P*=0.0476) (**Figure 3o**). Albeit statistically not significant, IFN-γ, IL-1RA and IL-6 levels were also elevated by trend in female Covid-19 patients with high SOFA scores compared to those with low SOFA scores (**Figure 3i-k**).

**Figure 3.**
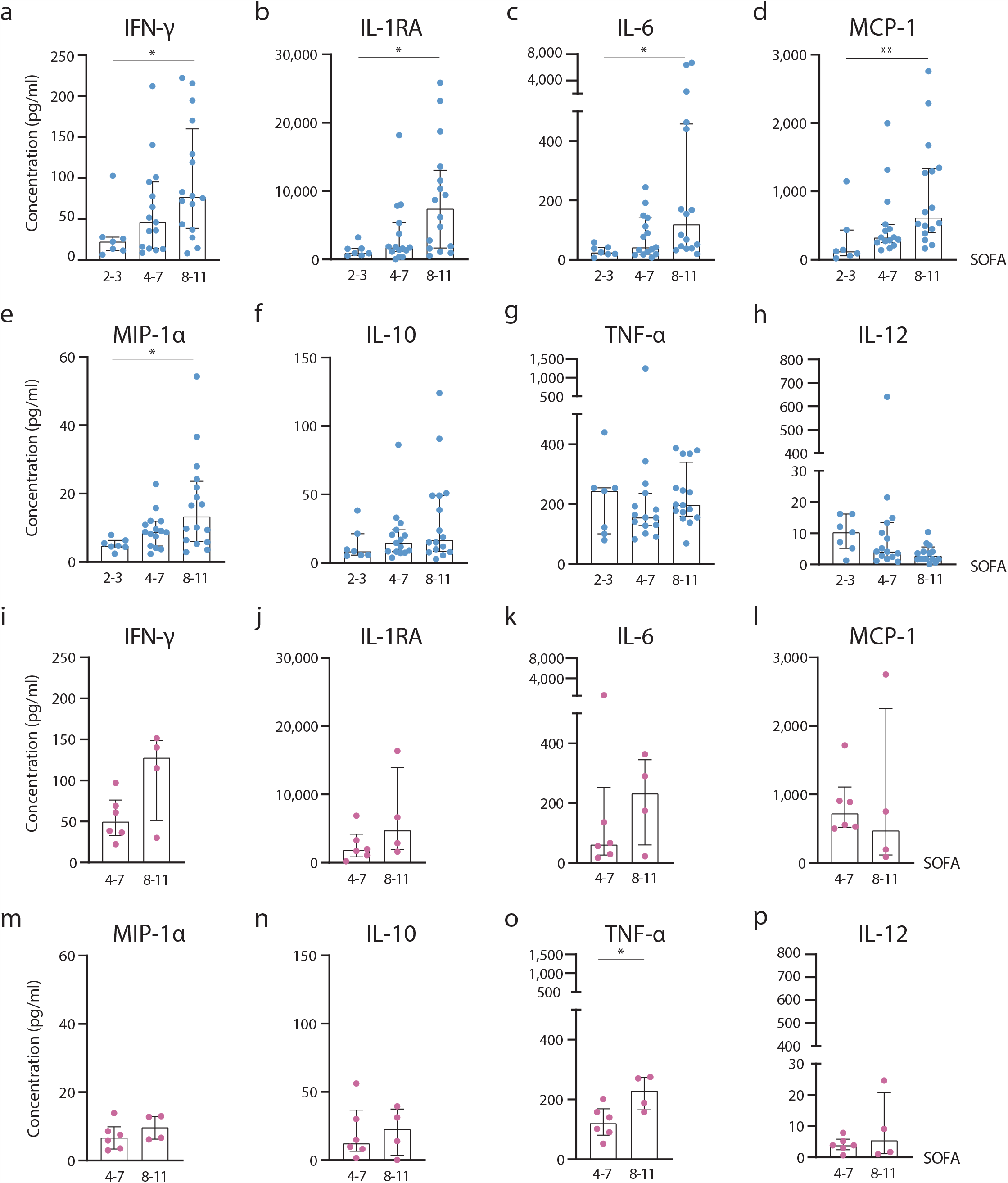
Chemokine and cytokine responses in Covid-19 patients. Shown are cytokine and chemokine levels of male and female COVID-19 patients in dependency of disease severity as assessed by SOFA scores (2-3; 4-7; 8-11). Blue dots represent males (SOFA 2-3, *n*=7; SOFA 4-7, *n*=15; SOFA 8-11, *n*=16) and pink dots represent females (SOFA 4-7, *n*=6; SOFA 8-11, *n*=4). Cytokine and chemokines were measured in plasma obtained from COVID-19 patients by using a 27-plex immunoassay. Here, those with significant differences are shown: IFN-γ (a,i), IL-1RA (b,j), IL-6 (c,k), MCP-1 (d,l), MIP-1α (e,m), IL-10 (f,n), TNF-α (g,o), IL-12 (h,p). Statistical significance in males was assessed by non-parametric tests (Kruskal-Wallis test and Dunn’s test for multiple comparisons). Statistical significance in females was evaluated by unpaired, two-tailed non-parametric Student’s *t*-test (Mann-Whitney test). Statistical significance was defined as *P*□<□0.05 (* *P*□<□0.05, ** *P*□<□0.01). Values are shown as median and interquartile range.

These findings show that cytokine and chemokine responses, particularly IFN-γ, IL-1RA, IL-6, MCP-1 and MIP-1α are generally elevated in dependency of disease severity.

### Sex hormone levels correlate with IFN-γ levels in Covid-19 patients

We next addressed the question whether changes in cytokine and chemokine responses in Covid-19 patients might correlate with their respective sex hormone levels given that most immune cells possess androgen and estrogen receptors ^9-11^. Performing linear regression analysis between all 27 cytokine and chemokines assessed, only IFN-γ presented a significant correlation to estradiol (R^2^=0.216, *P*=0.009; **Figure 4a**). Testosterone levels did not significantly correlate with changes in IFN-γ levels (R^2^=0.133, *P*=0.3111; **Figure 4b**).

**Figure 4.**
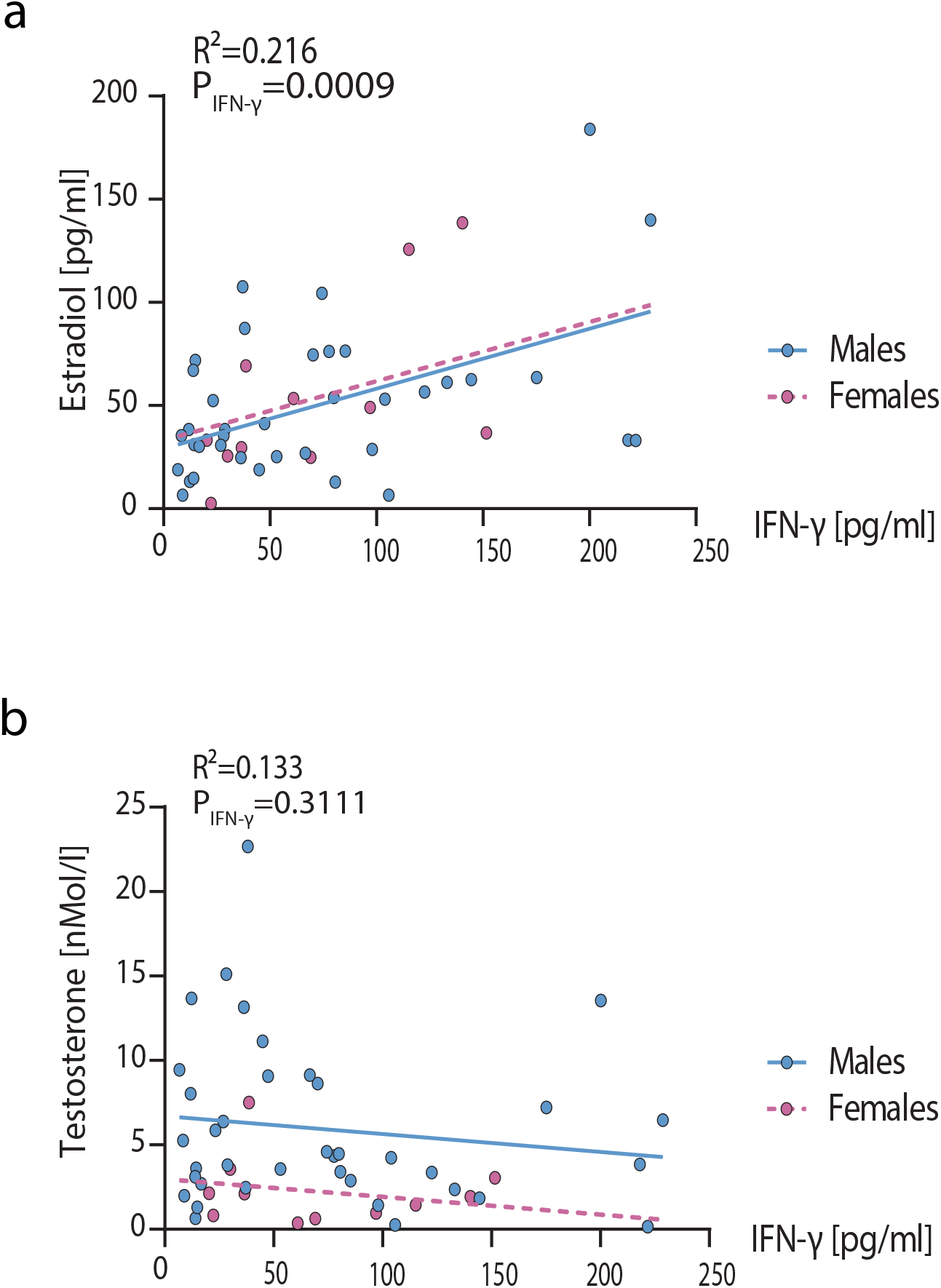
Correlation of IFN-γ levels in male and female COVID-19 patients to sex hormone levels. Testosterone and estrogen levels were measured in plasma of COVID-19 male (*n*=39) and female (*n*=11) patients and were plotted over the expression levels of all assessed cytokines and chemokines. Here, only IFN-γ is displayed, which showed significant correlations in regression analysis with estradiol (a) but not testosterone (b) levels among 27 different cytokines and chemokines assessed. Statistical significance was assessed by generalized linear regression.

These findings are in line with the estradiol-controlled transcription of IFN-γ since it possesses an estrogen responsive element (ERE) in its promoter region ^12-14^. IFN-γ is a key activator of macrophages ^14,15^ and macrophage activation was repeatedly reported as a hallmark of Covid-19 severity ^16^.

### Estradiol levels are associated with disease severity in male Covid-19 patients

Next, we analyzed whether sex hormone levels correlate with an increased risk for severe disease outcome as assessed by SOFA scores or the requirement for extracorporeal membrane oxygenation (ECMO) later during their ICU stay. Estradiol levels were elevated with increasing disease severity in male Covid-19 patients (*P*=0.0245 and *P*=0.0273) (**Figure 5a**). In female Covid-19 patients, estradiol levels also slightly increased with increasing disease severity, albeit statistically not significant likely due to the low sample size (**Figure 5b**). Male Covid-19 patients requiring ECMO treatment later during their ICU stay presented statistical significant higher estradiol levels than those not requiring ECMO during their later stay (*P*=0.0307) (**Figure 5c**). Testosterone levels did not show statistical significant changes comparing groups with different disease severity or requiring ECMO treatment in male or female patients (**Figure 5d-f**). In males, this is likely due to the fact that most male patients presented low testosterone levels below clinical references ^7,8^. Within the female Covid-19 cohort only 1 patient required ECMO treatment; thus, not allowing statistical analysis. As an additional parameter for disease severity, we analyzed endothelial lipase levels (EL) as a marker for endothelial activation. EL is a phospholipid hydrolyzing plasma lipase that is secreted by vascular endothelial cells and is involved in adhesion of monocytes to the endothelial cell surface under inflammatory conditions ^17^ and thus contributing to endothelial inflammation ^18^. In both, male and female Covid-19 patients, EL levels were significantly elevated compared to healthy controls (both *P*<0.0001) (**Figure 5g and h**). However, EL levels did not correlate with testosterone or estradiol levels (data not shown). Interestingly, levels of adiponectin, which is involved in the inhibition of EL secretion from activated endothelial cells ^18^, were not altered in male or female Covid-19 patients compared to the healthy cohort (**Figure 5 i and j**).

**Figure 5.**
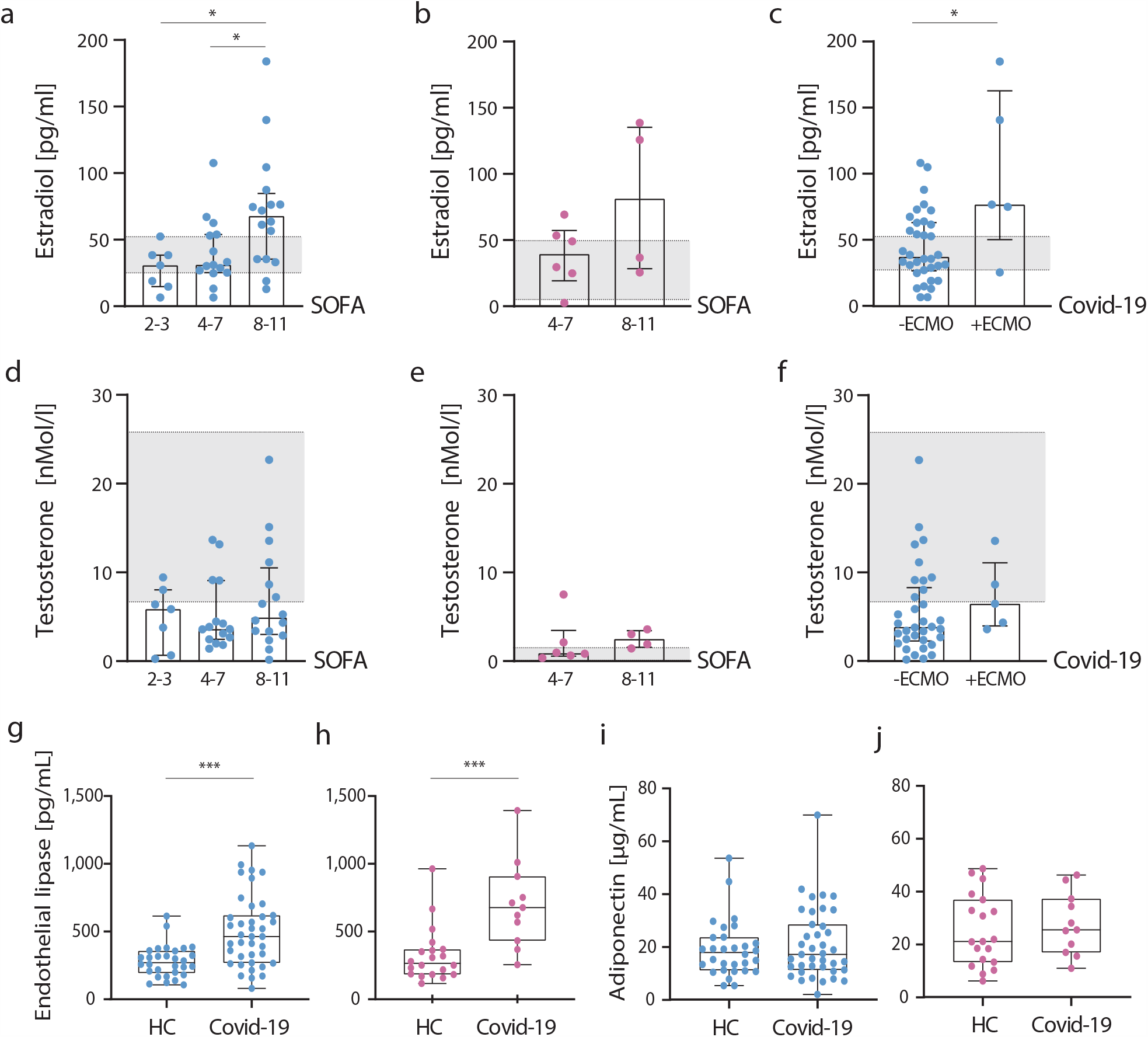
Sex hormone levels in COVID-19 patients in dependency of disease severity. Testosterone (a-c) and estradiol (d-f) levels were measured in plasma obtained from COVID-19 patients and are displayed in dependency of disease severity as assessed by SOFA scores (2-3; 4-7; 8-11). Blue dots represent males (SOFA 2-3, *n*=7; SOFA 4-7, *n*=15; SOFA 8-11, *n*=16) and pink dots represent females (SOFA 4-7, *n*=6; SOFA 8-11, *n*=4). Male COVID-19 patients were further subdivided into patients requiring connection to an ECMO (+ECMO, *n*=5) and patients not being placed on ECMO (-ECMO, n=34) (c,f). The laboratory assessed hormone reference ranges are indicated in grey. Statistical significance was assessed by Student’s *t*-test (* P□<□0.05, *** P□<□0.001). Endothelial lipase (g,h) and adiponectin (i,j) levels were measured in the plasma from COVID-19 patients and aged-matched healthy donors. Blue dots represent males (HD, *n*=30; COVID-19, *n*=30) and red dots represent females (HD, *n*=20; COVID-19, *n*=11). Values are shown as median. Statistical significance was assessed via Two-Way-ANOVA.

These data suggest that increased levels of estradiol and EL are associated with Covid-19 in both sexes.

### Metabolic profiling of Covid-19 patients reveals triglyceride depletion in males

Next, we wanted to study whether Covid-19 is also associated with alterations in metabolite profiles ^19^ besides the herein shown changes in steroid metabolism. Therefore, we measured 630 metabolites from 26 biochemical classes in male and female Covid-19 patients compared to healthy controls. Bioinformatics revealed distinct metabolite signatures in males and females, separating male from female in an unsupervised PCA analysis (**Figure 6a**). Mainly, significantly differentially regulated metabolites were identified in both directions in male compared to female Covid-19 patients (**Figure 6b and c**). These involved various pathways, such as primary bile acid biosynthesis, linoleic acid metabolism and glycerophospholipid metabolism (**Figure 6d**). However, in male patients, metabolites involved in primary bile acid biosynthesis and taurine and hypotaurine metabolism were up-regulated compared to female patients (**Figure 6e**). In contrast, metabolites involved in linoleic acid metabolism, alpha-linoleic acid metabolism, glycerolipid metabolism and sphingolipid metabolism are strongly down-regulated in male compared to female Covid-19 patients (**Figure 6f**). Herein, particularly 14 of the overall 21 down-regulated metabolites in male Covid-19 patients were triglycerides (**Figure 6g**).

**Figure 6.**
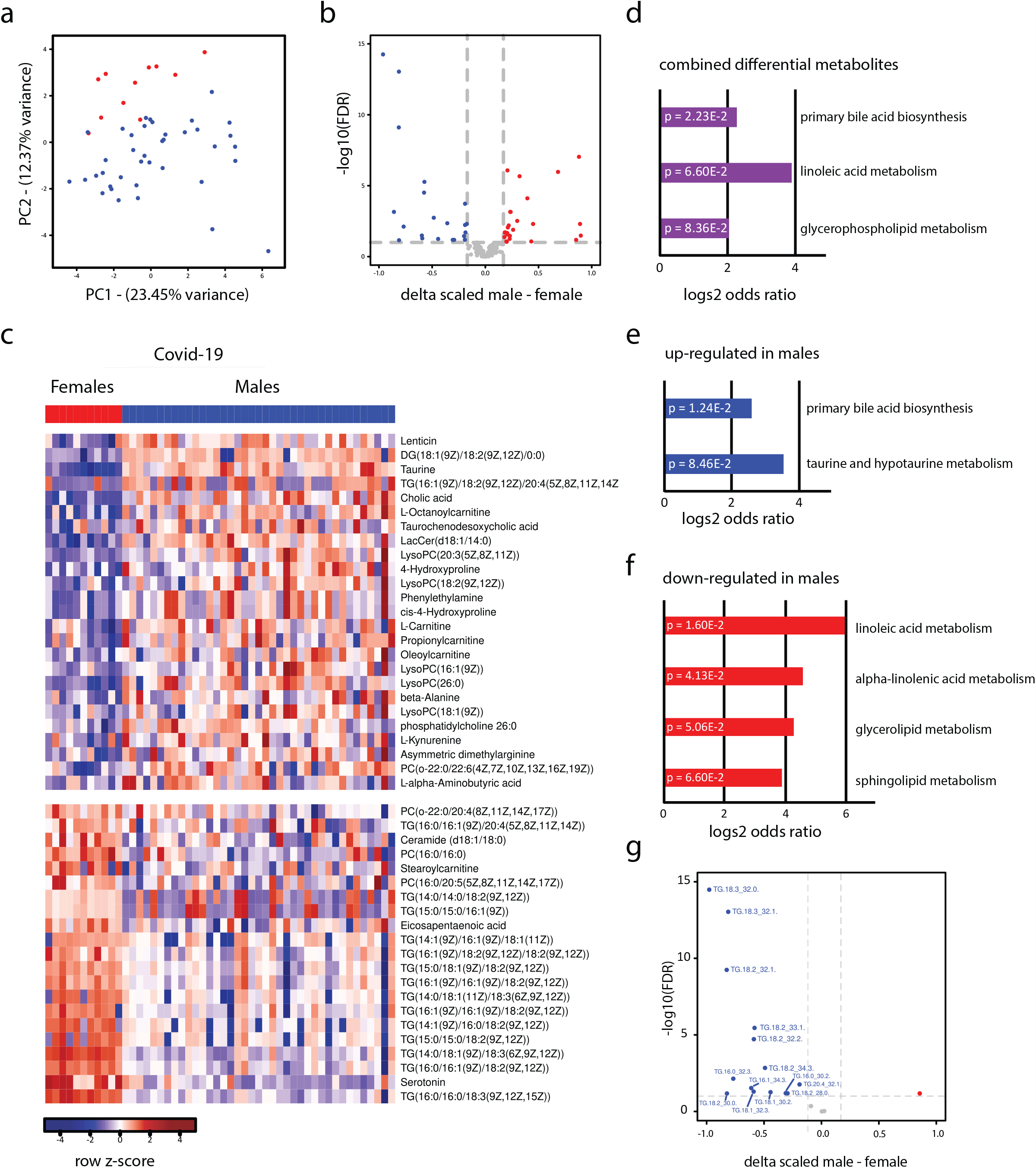
Metabolome profiling in male and female Covid-19 patients. PCA analysis (a) of metabolome profiles for male (blue; n=39) and female (red; n=11) Covid-19 patients using scaled normalized values (after subtracting gender-specific healthy control metabolome profiles). Differential analysis (b,c) between male and female Covid-19 patients; unpaired two-sided t-test followed by multiple testing correction was performed. Significant outliers are highlighted as blue or red in (b) or shown in (c) if FDR < 0.1 and delta male –female < -0.1699 or delta male –female > 0.1699 (reflecting a 12.5% difference). Pathway analysis was performed for all differential metabolites (d), only upregulated in male versus female (e) or only downregulated in male versus female (f); hypergeometric test was performed against the KEGG metabolite database ^32^ and all pathways with P<0.1 are shown. Differential analysis showing specifically all analyzed triglycerides (g); analysis performed as in (b).

These findings reveal strongly reduced triglyceride levels in male Covid-19 patients compared to female Covid-19 patients as a unique metabolic signature.

## Discussion

Metabolic and steroid hormone analysis of critically-ill Covid-19 patients in comparison to age- and sex-matched patients with coronary heart diseases (CHD) -as a male-biased top comorbidity in our Covid-19 cohort-as well as healthy controls revealed unique metabolic signatures.

First, we detected severely reduced testosterone levels in Covid-19 males compared to HC or CHD patients. Estradiol levels were not changed in male healthy controls or male CHD patients analyzed herein but were strongly elevated in Covid-19 males. The vast majority (95%) of testosterone is produced in Leydig cells of the testes depending on stimulation by luteinizing hormone (LH). Only small amounts (5%) are produced in the adrenal glands. Low levels of testosterone may either be of testicular origin (primary hypogonadism), of hypothalamic-pituitary origin (secondary hypogonadism) or a combination of both, which is predominantly found in the aging male population as late onset hypogonadism ^20,21^. Hypogonadism with and without elevated estradiol levels was reported before in patients with cardiovascular diseases as a risk factor for increased mortality in men ^6,22,23^. Thus, extrapolating from these reports on hypogonadism in males with cardiovascular diseases, the more severely reduced testosterone levels in the male Covid-19 cohort identified herein, further highlights the high risk for males. Furthermore, it is tempting to speculate whether an initial comorbidity-driven hit with respect to low testosterone levels (also reported for patients with obesity and type II diabetes ^24,25^, all top Covid-19 comorbidities) might put males at higher risk to develop severe Covid-19. This hypothesis is strengthened by recent findings in the golden hamster model showing that SARS-CoV-2 replicates in the reproductive system (testes, ovaries and uterus) of male and female animals. As a result, male animals present reduced testosterone and elevated estradiol levels ^4^. This is fully in line with our findings in the Covid-19 male cohort, which present reduced testosterone levels combined with elevated estradiol levels. Whether this is due to increased aromatase CYP19A1 (converts testosterone-to-estradiol) mRNA levels in the lung as suggested in the pre-clinical animal model ^4^, is unclear and requires further investigation. Noteworthy, the herein identified reduced testosterone levels were of testicular origin only in 25% of all male Covid-19 cases. Thus, future investigations regarding the impact of non-gonadal organs, such as the lung, in testosterone-to-estradiol aromatization in Covid-19 patients are required. We furthermore observed that some female Covid-19 patients also presented elevated testosterone levels albeit statistically not significant. In our female Covid-19 cohort, all except one patient, were postmenopausal. However, the low female Covid-19 cohort size in our study is a potential limitation with respect to conclusions on female Covid-19 outcome. This was due to the fact that more men than women were admitted to the ICU in the time period of recruitment further highlighting the importance of sex on critical Covid-19 outcome. Despite these limitations in the female Covid-19 cohort, other postulated an elevated Covid-19 risk for women with polycystic ovary syndrome (PCOS), a condition characterized by increased androgen levels ^26^. This highlights the need for further investigations to understand the impact of elevated testosterone levels in women in the context of Covid-19.

Second, we found that similar to males, female Covid-19 patients also present elevated estradiol levels. This is in contrast to the findings in the female SARS-CoV-2 golden hamster model using young, lean and comorbidity-free animals ^4^. Thus, future in depth investigations are required to understand the role of estradiol in postmenopausal Covid-19 patients. One possible investigation line might be to analyze whether elevated female testosterone levels might provide a substrate for the CYP19A1 aromatase resulting in elevated estradiol levels in at-risk postmenopausal women upon SARS-CoV-2 infection. However, elevated estradiol levels seem to be a risk factor for severe Covid-19 outcome in male and female patients. Male Covid-19 patients on the other hand, additionally suffer from severely depleted testosterone and dihydrotestosterone levels, which might present an additional hit putting them at increased risk compared to females.

Third, by analyzing 27 different cytokines/chemokines and correlating their levels to testosterone or estradiol levels using regression analysis, we identified interferon-γ (IFN-γ) as a key cytokine that positively correlated with estradiol levels. This finding is of highest importance given that IFN-γ is the key cytokine responsible for macrophage activation ^14,15^. IFN-γ primes macrophages that are activated by external TLR stimuli, such as viral infections and then secrete higher level of pro-inflammatory but lower level of anti-inflammatory cytokines ^27^. Macrophage activation in the lung upon SARS-CoV-2 infection was repeatedly reported to be a hallmark of fatal Covid-19 outcome ^16^. Macrophages contain membrane-bound as well as nuclear androgen- and estrogen-receptors. Thus, it will be of high interest to dissect the role of sex hormones and IFN-γ in orchestrating e.g. macrophage activation, cytokine storm and endothelial activation pathways in future studies (**Figure S1**).

Fourth, analyzing over 600 metabolites representing various biochemical pathways, we identified lower triglycerides as a unique feature in critically-ill Covid-19 patients. Triglycerides are usually elevated in patients with sepsis. Inflammation-related inhibition of lipoprotein lipase due to hyperglycaemia and hyperinsulinaemia produces upregulated hepatic triglyceride production. In addition, disturbances of the mitochondrial fatty acid β-oxidation also lead to higher levels of triglycerides ^28^. The underlying cause of reduced triglycerides and altered bile acid profiles in male Covid-19 males is not clear but indicate an involvement of the liver, which is the responsible organ for the secretion of triglyceride-rich lipoproteins and bile acids ^29^. In future, the disturbed lipid metabolism need particular attention given that comorbidities, such as obesity are usually associated with elevated triglyceride levels ^24^. Interestingly, some of these species have a key role in regulating macrophage functions ^30^, as lipids are not only a source of energy for macrophages but they are especially important to provide precursors for bioactive lipids. Altered plasma concentrations of those lipid species indicate a dysregulated lipid metabolism that could impact macrophage functions and thus severity of infectious diseases including Covid-19 ^30^. Thus, future studies are needed to understand the complex cross-talk between lipid metabolism, endothelial cell and macrophage activation in male Covid-19 patients.

Collectively, our findings herein highlight that metabolic dysregulations pose a hallmark of severe Covid-19 outcome. In particular, monitoring sex hormone and triglyceride levels might offer new diagnostic opportunities for patient management and mitigation of early intervention strategies.

**Table S1:**
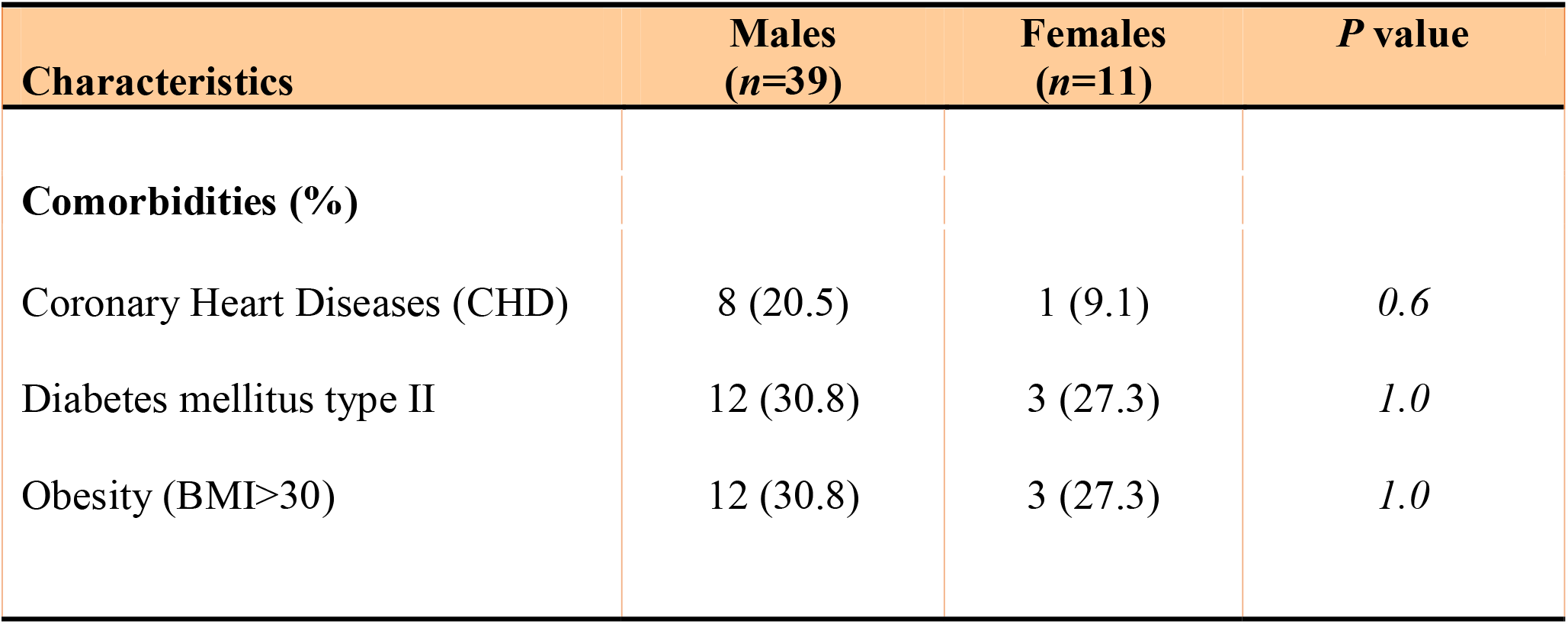
Covid-19 Comorbidities.

| Characteristics | Males<br>(*n*=39) | Females<br>(*n*=11) | *P* value |
| --- | --- | --- | --- |
| **Comorbidities (%)** |  |  |  |
| Coronary Heart Diseases (CHD) | 8 (20.5) | 1 (9.1) | *0.6* |
| Diabetes mellitus type II | 12 (30.8) | 3 (27.3) | *1.0* |
| Obesity (BMI>30) | 12 (30.8) | 3 (27.3) | *1.0* |

## Methods

### Ethics statement

Sampling from laboratory-confirmed Covid-19 patients was reviewed and approved by the ethics committee at the Hamburg State Chamber of Physicians (registration no.: WF-053/20). The need for an informed consent for healthy blood donors (HC) and patients with coronary heart diseases (CHD) was waived by the ethics committee because data were retrieved retrospectively from electronic health records.

### Study design, setting and participants

This retrospective cohort study included plasma samples collected from the first 50 laboratory-confirmed COVID-19 patients who were admitted to the ICU at the University Medical Center Hamburg-Eppendorf from March 8^th^ to April 29^th^, 2020. A total of 39 male and 11 female patients were included in this study. The University Medical Center Hamburg-Eppendorf is a tertiary care hospital with 1,738 hospital beds. The Department of Intensive Care Medicine includes 12 multidisciplinary ICUs with a total of 140 ICU beds, regularly. During the SARS CoV-2 pandemic, 3 intensive care units were initially designated as dedicated units for the treatment of COVID-19 patients. Up to 30 patients suffering from COVID-19 are currently being treated here. A further ward treated critically-ill patients with suspected COVID-19 until laboratory confirmation of SARS-CoV-2. The in-house crisis management team has developed a step-by-step plan for this purpose. Normally, the Department of Intensive Care Medicine has units for the care of all specialties, including specialized units for the treatment of acute respiratory distress syndrome (ARDS), which routinely use extracorporeal membrane oxygenation (ECMO). All samples, including plasma samples from 34 male and 11 female patients as well as serum samples from 5 male and one female patients, were collected from COVID-19 patients on the day of admission (except one sample, which was collected 1 day later) and stored at -80°C until further analyses. Methods were validated for serum samples by analyzing plasma and serum samples obtained from the same patients. Plasma samples from healthy donors were collected from the blood donation center of the transfusion medicine at the University Medical Center Hamburg-Eppendorf. Coronary heart disease patient plasma samples were collected at the University Hospital in Tübingen.

### Data collection

The following demographic and clinical variables were collected retrospectively from the electronic patient data management system (PDMS) (ICM, Dräger, Lübeck, Germany): age, sex, body mass index, comorbidities, Simplified Acute Physiology Score II (SAPS II) on admission, Sequential Organ Failure Assessment Score (SOFA) on admission, and classification of Acute Respiratory Distress Syndrome (ARDS) using the Berlin definition^9^. Additionally, we recorded antiviral treatment, supportive and experimental COVID-19 therapies, the need for mechanical ventilation and extracorporeal membrane oxygenation^10^. Furthermore, we followed the course of the patients and recorded discharge or death.

### Hormone quantification

A panel of 13 hormones was measured in plasma samples of COVID-19 patients (total testosterone, free testosterone, dihydrotestosterone, androstenedione, 17-β-estradiol, estrone, sex hormone-binding globulin, thyroid-stimulating hormone, free triiodothyronine (T3), free thyroxine (T4), luteinizing hormone, follicle-stimulating hormone and cortisol), as well as testosterone, dihydrotestosterone, estradiol and estrone levels of healthy donor samples, by an external laboratory accredited for measurements of human samples (Labor Lademannbogen, Hamburg, Germany). Cortisol, TSH, T4, LH, FSH, TT, E2 and SHBG were analyzed by electro-chemiluminescence immunoassay (ECLIA). Free TT was analyzed by enzyme-linked immunosorbent assay and DHY-TT was measured by liquid chromatography–mass spectrometry (LC-MS/MS). Estrone levels were measured by a radioimmunoassay (RIA).

Testosterone plasma levels of coronary heart disease patients were measured in a custom-made MILLIPLEX MAP Multi-Species Hormone Magnetic Bead Panel (Merck), according to the manufacturer’s instructions in a Bio-Plex 200 System with high-throughput fluidics (HTF; Bio-Rad). Estradiol (Hölzel Diagnostika) plasma levels of coronary heart disease patients were analyzed by ELISA following the manufacturer’s instructions and measured on an ELISA microplate reader (Saphire2, Tecan).

### Measurement of adiponectin and endothelial lipase levels

Adiponectin (Sigma-Aldrich) and endothelial lipase (IBL international) levels in plasma samples were evaluated by ELISA following the manufacturer’s instructions. For adiponectin analysis, samples were diluted 1:500. All ELISAs were measured on a Saphire2 ELISA microplate reader (Tecan) and evaluated using a four parameter logistic regression (MyAssays).

### Cytokine and chemokine measurement

A panel of 27 cytokines and chemokines (eotaxin, fibroblast growth factor (FGF), granulocyte-colony stimulating factor (G-CSF), interferon-γ (IFN-γ), interferon γ-induced protein (IP-10), interleukin-2 (IL-2), interleukin-4 (IL-4), interleukin-5 (IL-5), interleukin-6 (IL-6), interleukin-7 (IL-7), interleukin-8 (IL-8), interleukin-9 (IL-9), interleukin-10 (IL-10), interleukin-12 (IL-12), interleukin-13 (IL-13), interleukin-15 (IL-15), interleukin-17 (IL-17), interleukin-1β (IL-1β), interleukin 1 receptor antagonist (IL-1RA), monocyte chemoattractant protein-1 (MCP-1), platelet-derived growth factor-BB (PDGF-BB), regulated upon activation, normal T-cell expressed and presumably secreted chemokine (RANTES), tumor necrosis factor-α (TNF-α), and vascular endothelial growth factor (VEGF)) was measured in plasma samples of COVID-19 patients. Cytokine and chemokine levels were measured using a Bio-Plex Pro™ multiplex assay (#M500KCAF0Y, Bio-Rad, Feldkirchen, Germany) according to the manufacturer’s instructions in a Bio-Plex 200 System with high-throughput fluidics (HTF; Bio-Rad, Feldkirchen, Germany).

### Metabolic Profiling

Before shipment, serum samples were inactivated by adding ethanol at a sample-solvent ratio of 1:2 (v:v). For sample processing the MxP® Quant 500 Kit (BIOCRATES Life Sciences AG, Innsbruck, Austria) was applied following the protocol of the manufacturer. Briefly, 30 µL of inactivated serum sample, 10 µL of each calibration standard (n = 7) and control sample (n = 7) were transferred onto a filter containing internal standards for internal standard calibration. The filters were dried under a stream of nitrogen using a pressure manifold (Waters, Eschborn, Germany). Afterwards, the derivatization reagent phenyl isocyanate was added to each sample and incubated for 60 min. After drying under nitrogen, analytes were extracted with 5 mmol/L ammonium acetate in pure methanol and the eluate was further diluted for the LC-MS/MS analysis. The targeted analysis covered 630 metabolites from several biochemical classes (amino acids, amino acid metabolites, cholesterol esters, bile acids, biogenic amines, carboxylic acids, fatty acid, acylcarnitines, glycerophospholipids, sphingolipids, and triglycerides), which were detected by tandem mass spectrometry (MS/MS) after ultra-high pressure liquid chromatographic (UHPLC) separation and flow injection analysis (FIA), respectively. Each sample measurement required two UHPLC runs and three FIA runs to cover all metabolites. All analyses were performed on an UPLC system (ACQUITY UPLC I-Class, Waters) coupled to a triple quadrupole mass spectrometer (Xevo TQ-S, Waters). Chromatographic separation was accomplished using a reversed phase column (C18, BIOCRATES) with 0.2 % formic acid in water (solvent A) and 0.2 % formic acid in acetonitrile (solvent B) as eluent system. The gradient profile was in accordance with the protocol of the manufacturer. FIA solvent was methanol with a modifier, which was provided by the kit manufacturer. Data analysis of the UHPLC results was based on a seven-point curve or one-point calibration and internal standard normalization.

The bioinformatics analysis of the raw metabolite quantifications was done mostly in concordance with the Metaboanalyst pipeline ^31^ and performed in R version 4.0.2. First, missing values were replaced with minimum inferred values. Then, filtering was performed considering quality control samples with the relative standard deviation option with a cut-off of 25. Normalization of quantifications was performed using quantile-normalization followed by log-transformation. In order to eliminate any gender-specific variation of baseline metabolite levels between male and female, we first subtracted normalized log-transformed average metabolite values of either female or male healthy controls from each respective female or male Covid-19 sample and metabolite; these values are referred to as “scaled” throughout this article. Differential analysis between male and female Covid-19 samples was done using the aforementioned scaled values, applying an unpaired two-sided t-test followed by multiple testing correction (false-discovery rate or FDR). Pathway analysis was done using the KEGG database as provided by the Metaboanalyst implementation, using a hypergeometric test.

### Statistical analysis

Statistical evaluation for quantitative data was performed with two-way-ANOVA including the cohort and sex as independent variable as well as its interaction. For non-normal data unpaired Mann-Whitney or Kruskal-Wallis test ignoring any multiple comparisons were used. Statistical significance was defined as P□<□0.05 (* P□<□0.05, ** P□<□0.01 and *** P□<□0.001. All statistical evaluations mentioned above were performed with SAS®, version 9.4 TS level 1M5 (SAS Institute Inc., Cary, NC, United States). Graphical representation of the data was performed via GraphPad Prism 8 v. 8.4.2 (GraphPad Software, Inc.).

## Data Availability

All generated data are available in this study. No external repositories were used.

## Author contributions

GG conceived the study. SK, MS, DJ and AN were responsible for COVID-19 patient management and recruitment. GG, SK, MS and BS (Berfin Schaumburg) designed and overviewed the study design. BS, HJ, MZ and SSB conducted the cytokine and chemokine assays. ZK and BS performed hormone assays. MP performed the metabolome analysis. GG, BS, MS, AK and TB analyzed data and developed the figures. LK, BetS (Bettina Schneider) and FS conducted ANOVA models and statistical tests. JA, AM, TB, JH, HS critically reviewed the manuscript and were involved in study design. GG wrote the manuscript. All authors revised the manuscript.

## Acknowledgment

This study was supported by a rapid response grant from the Federal Ministry of Health (BMG) to G.G.

## Role of funding source

The study funder had no role in study design, data collection, data analysis, data interpretation, or writing of the report. The corresponding author had full access to all the data in the study and had final responsibility for the decision to submit for publication. MS receives funding from the Medical Faculty of the University of Hamburg for clinical leave.

## Declaration of Interests

All authors declare no competing interests.

## Figure Legends

**Figure S1.**
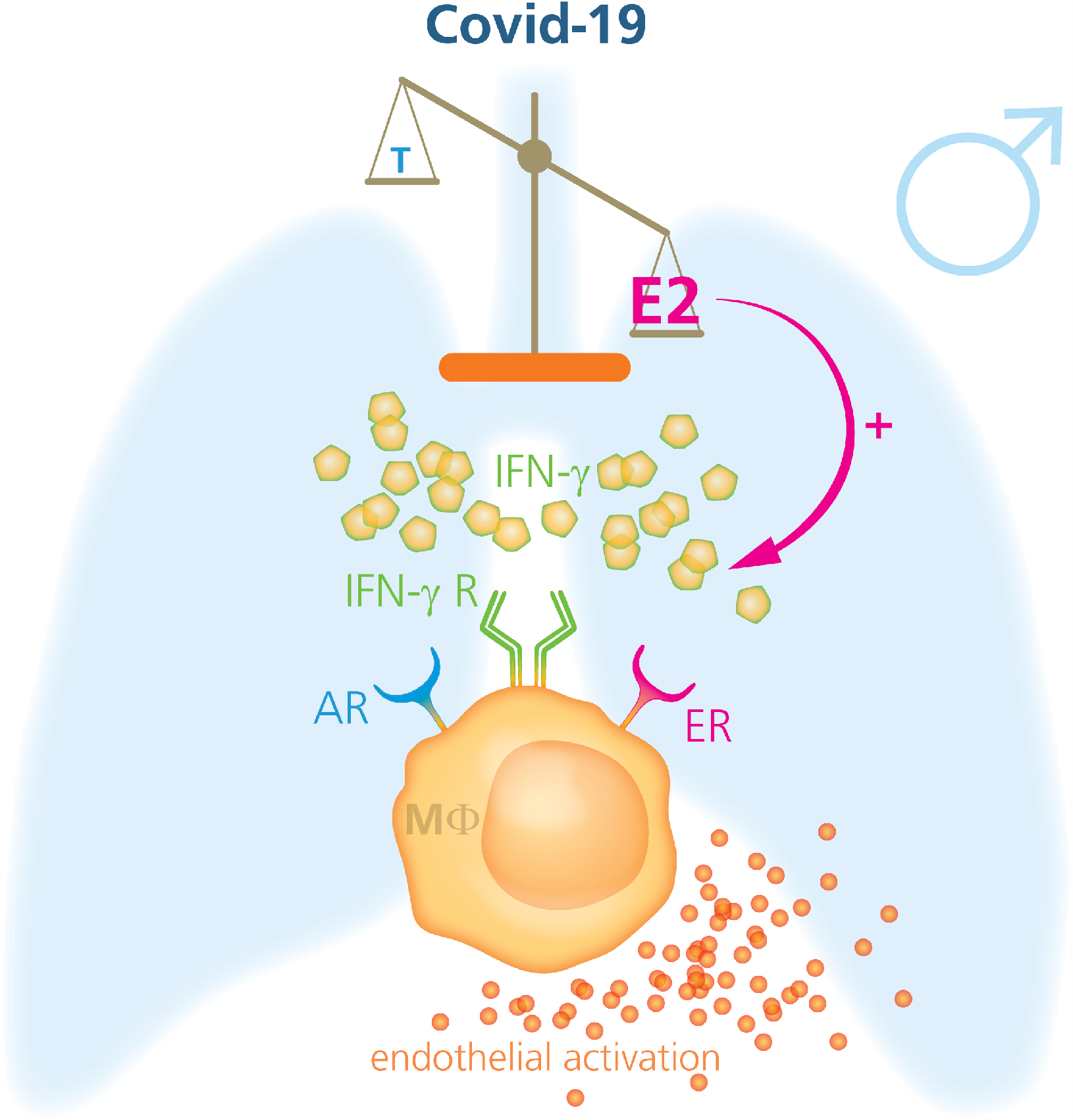
Model: Sex hormones triggered activation of immune pathways in Covid-19 males. Critically-ill men diagnosed with Covid-19 present reduced testosterone and increased estradiol levels. Regression analysis revealed that among 27 cytokines and chemokines analyzed, only IFN-γ levels showed significant correlation to sex hormone levels. Estradiol levels positively associated with IFN-γ levels. IFN-γ was repeatedly reported before to have an estrogen responsive element (ERE) in its promoter region ^12,13,33^ further highlighting the role of estradiol in particular in regulating IFN-γ transcription. Macrophages contain membrane-bound as well as nuclear acting androgen and estrogen receptors. Macrophages also contain an IFN-γ receptor. IFN-γ poses a major cytokine that primes macrophages for the secretion of inflammatory cytokines upon external TLR stimuli, such as viral infections ^27^. Macrophage activation was proposed to play a key role in Covid-19 pathogenesis ^14-16^. We further identified elevated endothelial lipase levels as a potential marker of endothelial activation in Covid-19 patients. Collectively, we herein propose that sex hormones and IFN-γ play key roles in macrophage-mediated activation and endothelial damage in Covid-19 patients.

## Notes

### Competing Interest Statement

The authors have declared no competing interest.

### Funding Statement

This study was funded by the German Ministry of Health.

